# Association between Dental Caries and Unemployment among U.S. Adults with a history of Illicit Drugs

**DOI:** 10.1101/2025.08.01.25332750

**Authors:** Sucharu Ghosh, Samarpita Chatterjee, Changyong Feng, Janine Burkhardt, Sangeeta Gajendra

## Abstract

**Objectives:** This study aims to explore the relationship between dental caries and unemployment among U.S. adults who have engaged in illicit drug use, such as cocaine, heroin, and methamphetamine.

**Methods:** The National Health and Nutrition Examination Survey (2015-2018) data were analyzed. The independent variable was severe dental caries (defined as DMFT >13.99), and the dependent variable was employment status. The sample was categorized into non-users, current users (used in the past year), and former users (used prior to the past year). Covariates included age, education, race, gender, smoking status, family income-to-federal poverty level ratio, and health insurance status. Logistic regression with survey weights was applied to assess associations between severe dental caries and employment status.

**Results:** The total sample (n=5,476) represented 131,848,604 U.S. adults aged 18-59 years, with 4% current users and 12% former users of the specified drugs. Among current users, those with severe caries had higher odds of unemployment (OR = 2.6, p = 0.025) compared to those without severe caries. No significant association was found between severe caries and employment status among former users after controlling for covariates.

**Conclusions:** The study underscores a significant association between severe dental caries and unemployment among U.S. adults who have used illicit drugs in the past year. These findings suggest a potential need for targeted oral health interventions in this population to improve economic well-being. Future research should focus on longitudinal studies to establish causality and explore mechanisms through which dental health may impact employment prospects.

## Background and Literature Review

The use of illicit substances, including cocaine, heroin, and methamphetamine, poses a significant public health concern. While substance use often begins voluntarily, addiction creates barriers to cessation and presents complex challenges for public health interventions (1).

Social determinants of health impact an individual’s ability to maintain health within their community, including factors such as neighborhood, access to healthy food, housing, education, and economic stability (2). The social-ecological model (SEM) elucidates primary causes of health inequalities at individual, interpersonal, community, and societal levels (Fig. 1) (3). On an individual level, factors like physical and mental health, withdrawal management, harm reduction knowledge, and employment status affect illicit drug use behaviors. Interpersonal influences include access to drugs, attitudes towards substance use, peer and family support, and acceptance of medication-assisted treatment (MAT). At the community level, culturally competent provider access, behavioral health services, and drug disposal facilities play a role. Societal factors encompass stigma towards drug users, policies promoting equity, health insurance coverage, and housing stability. Effective interventions often operate across multiple SEM levels (3).

**Fig. 1:**
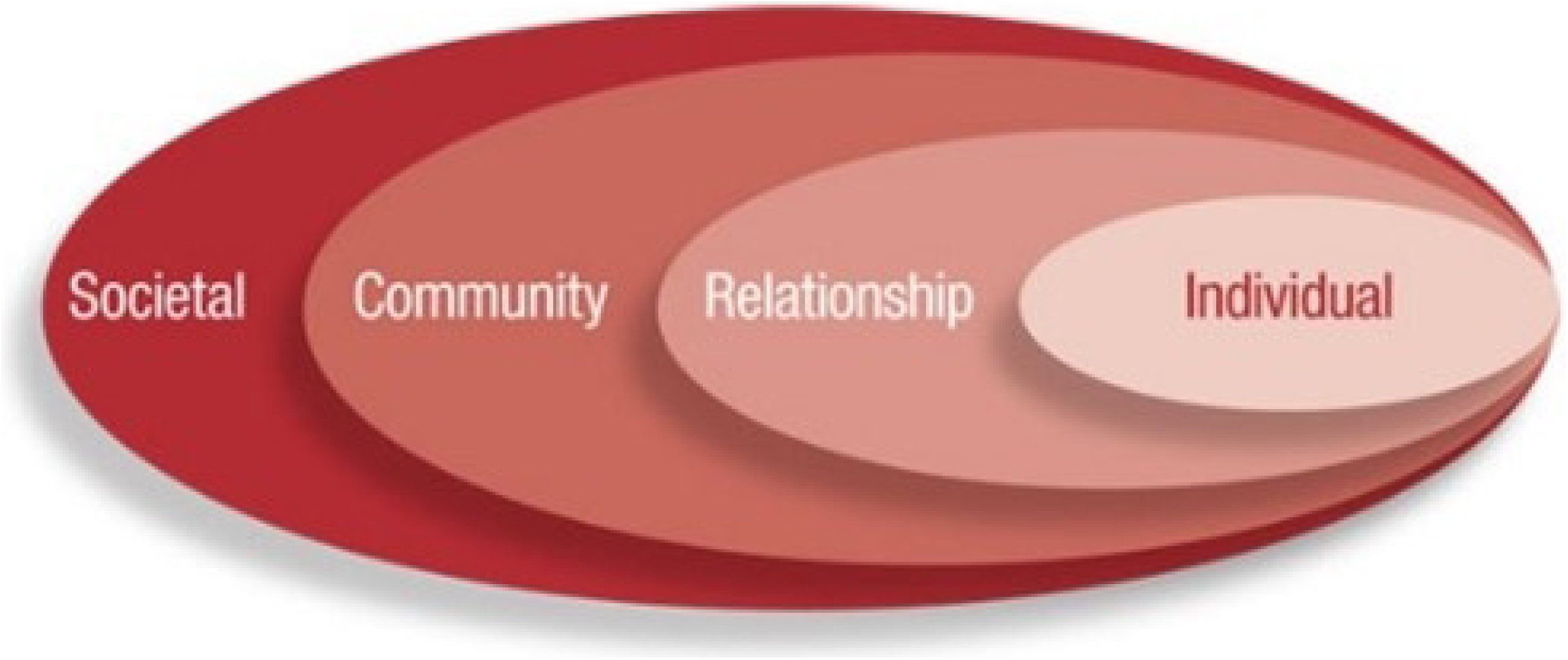
The socioecological model

**Fig. 2:**
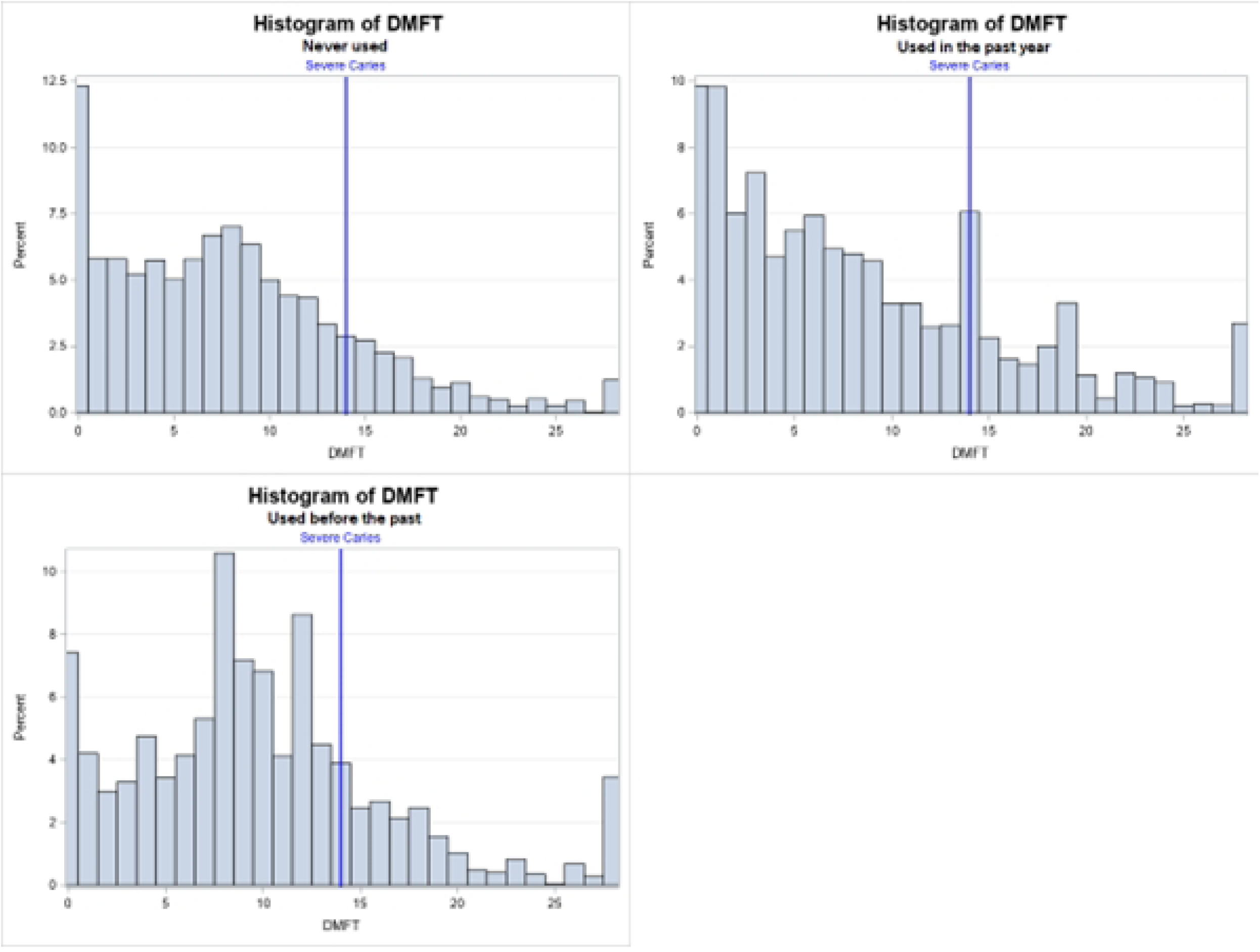
Histogram of DMFT stratified by a) never used, b) used in the past year, and c) used before the past year.

Socioeconomic status, education, healthcare access, and housing stability critically influence both oral health and employment prospects. Socioeconomic disparities can limit oral health access and outcomes (4), while education level affects substance use behaviors, health literacy, and employment opportunities (5). Access to healthcare is linked to employment status, influencing oral health care and maintenance (6). Though limited in research, housing stability also plays a significant role in substance abuse and oral health outcomes.

Illicit drug use has widespread public health implications, contributing to increased morbidity, mortality, and healthcare costs estimated at $193 billion annually in the U.S. Comorbidities, such as mental health disorders and infectious diseases, exacerbate the challenge of effective public health interventions (7, 8, 9).

Illicit drug use puts individuals at a greater risk for poor oral health (10). Limited or no access to dental care, poor oral hygiene maintenance, and poor dietary habits are direct adverse effects of substances on oral health. Inferior quality of diet and self-care lead to higher rates of dental caries, enamel erosion, and periodontal disease (11). These oral health issues not only affect physical health but also have significant psychosocial impacts, including negative effects on esthetics, self-esteem, and self-confidence (12). Furthermore, opioids, anti-depressants, and antipsychotics like phenothiazines used for the management of drug addiction are often associated with salivary hypofunction, which may lead to significantly higher prevalence of caries (13, 14). An analysis of the 2009-2014 National Health and Nutrition Examination Survey (NHANES) showed that current cocaine users had 38.5% higher untreated dental caries compared to non-users. The study also found that untreated dental caries was more likely to be found among cocaine users with cigarette smoking (OR=1.94; 95% CI=1.21-3.11) or with use of methamphetamine (OR=5.40; 95% CI=1.92-15.14) than only cocaine users (15). Hegazi et al (16) found higher untreated dental caries in current (PR: 1.53; 95% CI: 1.10-2.13) and established (PR: 1.21; 95% CI: 1.02-1.48) methamphetamine users compared to never users (10). While these studies were comprehensive, they did not explore the social determinants of health such as unemployment faced by illicit drug users.

Social determinants of health provide a crucial context for examining the relationship between dental caries, employment, and drug use. Unemployment not only impacts individual well-being but also contributes to broader economic issues, including increased healthcare costs and loss of productivity (17, 18).

The impact of dental health on employment status has been explored in several studies. Study by Al-Sudani et al. (19) and Halasa-Rappel et al. (20) independently found significant associations between unemployment and numbers of missing and decayed teeth. These studies were methodologically sound, using clinical assessments to determine oral health status, nationally representative sample and it emphasized the broader socioeconomic implications of poor oral health. However, they do not delve into the specific challenges faced by adults with a history of illicit drug use.

The relationship between substance use and employment status has been well-documented in the literature. The Substance Abuse and Mental Health Services Administration (SAMHSA) reported that 8.6 percent full-time workers in the U.S. used illicit drugs in the past month (21). Truong et al. found that employed people seldom experienced negative impacts due to their oral health than the people who were unemployed (22). Additionally, Hanson et al. (23) found a 460% increase in the employment rate among patients who received comprehensive oral health care versus only a 130% increase among those who did not in a treatment facility. Supic et al. found unemployment (OR=3.23) to be a significant predictor of poor oral health in heroin users (24). However, these studies often lack a specific focus on the oral health challenges of illicit drug users, leaving an important research gap.

Despite growing evidence that substance use can adversely impact oral health, few studies have explored how these oral health challenges intersect with employment outcomes among adults who use illicit drugs. Employment is a key social determinant of health, providing economic stability, social support, and access to health resources. By focusing on a nationally representative sample, our study contributes new insights into how illicit drug use could influence oral health and, ultimately, employability. This addresses a gap in the literature, underscoring the need for multidisciplinary prevention and intervention strategies that account for both socioeconomic factors and varying levels of substance use.

## Methods and procedures

### Study design

This cross-sectional study utilized data from the National Health and Nutrition Examination Survey (NHANES) from 2015 to 2018 (25). The survey uses a three-stage stratified random sampling method to sample about 5000 people each year that yields nationally representative data. The survey over-samples persons 60 and older, African Americans, and Hispanics to attain adequate numbers of participants in those groups to support sub-group comparisons. Data collection includes both interviews (demographic, socioeconomic, dietary, and health-related questions) and physical examinations (medical, dental, physiological measurements, and laboratory tests) conducted by trained professionals.

### Ethics Statement

NHANES 2015-18 was approved by The National Center for Health Statistics (NCHS) Research Ethics Review Board (26). This study was considered as an exempt study by the University Rochester Research Subjects Review Board.

### Study Population and Variables

Adults 18 years and older were included in this study. Based on the reported use of cocaine, heroin, and methamphetamine, the total population was divided into three groups. Participants who answered ‘No’ to the survey question, ‘Ever used cocaine/heroin/methamphetamine?’, were categorized under “Never used”. Those who used cocaine, heroin, or methamphetamine within the past year were categorized as “Current users” and those who used prior to the past year were considered as “Former users”.

The independent or exposure variable of the study was caries experience, which was measured by the decayed, missing, and filled teeth (DMFT) index Severe caries was defined as a DMFT score ≥13.99, based on the criteria established by Petersen et al. in the Bulletin of the World Health Organization (27). The DMFT index was calculated for each participant using examination data, excluding third molars. The following conditions were counted as decayed: “Permanent root tip is present, but no restorative replacement is present”, “Primary tooth with a dental carious surface condition”, and “Permanent tooth with a dental carious surface condition”. The following tooth conditions were counted as missing: “Missing due to dental caries”, “Missing due to dental caries but replaced by a removable restoration”, and “Missing due to dental caries but replaced by a fixed restoration”. Conditions that were counted as filled were: “Primary tooth with a restored surface condition”, “Permanent tooth with a restored surface condition”, and “Permanent root tip is present, but a restorative replacement is present”.

The primary outcome of our study was employment status. The survey question: ‘type of work done last week’ was used to measure the outcome variable. Participants who answered “Working at a job or business” or “with a job or business but not at work” were considered as employed. Both full-time and part-time employment was included. Participants who answered “looking for work” or “not working at a job or business” were coded as unemployed.

### Covariates and Confounders

The confounders were identified based on the literature review and directed acyclic graphs. Demographic variables such as age, education, race, gender, ratio of family income to the federal poverty level and health insurance as well as smoking status were included in the analyses. All covariates were directly available from the NHANES dataset. Age was a continuous variable but was categorized into the following five age groups: 18-30, 31-40, 41-50, 51-65. In the original dataset, education was coded as two separate variables for individuals less than 20 years of age and more than/equal to 20 years. To operationalize, these two variables were merged, and the new variable had the following categories: “less than 9th grade”, “9-11th grade (includes 12th grade with no diploma)”, “high school graduate or General Educational Development (GED) or equivalent”, “some college or associate degree”, and “college graduate or above”. Participants who reported that they smoked at least 100 cigarettes in their life were considered smokers.

### Statistical Methods

Pooled samples from the two NHANES cycles of 2015-2016 and 2017-2018 were used. Complete case analysis was performed and participants with missing data in any of the exposure, outcome, and covariates were excluded from the analysis.

Survey weights from NHANES were used to account for the complex sampling design, oversampling, non-response, and post-stratification adjustments to match U.S. Census data (28). Weights were recalibrated by dividing them in half for accurate representation. Descriptive analyses were conducted to assess demographic characteristics, mean DMFT, and prevalence estimates for dental caries across user groups.

Preliminary analyses indicated a significant association between severe caries and employment among current drug users. Multivariable logistic regression models were applied to estimate odds ratios (OR) and 95% confidence intervals (CI) for the association between severe caries and employment status among current users (Table 3). The four models included: (1) unadjusted (severe caries and employment status), (2) adjusted for variables associated with either the exposure or outcome, (3) further adjusted for variables that changed the log odds by ≥10%, and (4) fully adjusted for potential confounders. Model selection was based on at least a 10% change in OR. Analyses were performed using SAS 9.4, with statistical significance set at α=0.05 (29).

## Results

The study included 5,476 participants after excluding those with missing data, representing a weighted sample of 131,848,604 U.S. adults. Table 1 presents the distribution of demographic variables and dental caries status, stratified by drug use history: current users (used cocaine, heroin, or methamphetamine within the past year), former users (used drugs prior to the past year), and non-users (never used these drugs). Four percent of participants were current users, 12% were former users, and 84% were non-users.

**Table 1:** Table showing distribution of demographic variables and dental caries status stratified by people who used cocaine, heroin, or methamphetamine in the past year, used before past year, and never used (n=5476).

|  |  | Total | Never Used | Current user | Former User | p-value |
| --- | --- | --- | --- | --- | --- | --- |
|  |  | Weighted N (%) | N (%) | N (%) | N (%) |  |
|  |  | 131848604 (100%) | 4583 (84%) | 239 (4%) | 654 (12%) |  |
| **Age** | 18-30 | 36675280 (28%) | 1426 (31%) | 95 (40%) | 103 (16%) | <.0001 |
|  | 31-40 | 30730089 (23%) | 1027 (22%) | 69 (29%) | 178 (27%) |  |
|  | 41-50 | 31015312 (24%) | 1090 (24%) | 32 (13%) | 140 (21%) |  |
|  | 51-59 | 33427922 (25%) | 1040 (23%) | 43 (18%) | 233 (36%) |  |
| **Gender** | Male | 64761729 (49%) | 2070 (45%) | 166 (69%) | 380 (58%) | <.0001 |
|  | Female | 67086875 (51%) | 2513 (55%) | 73 (31%) | 274 (42%) |  |
| **Race** | Mexican/Other Hispanic | 22347937 (17%) | 1245 (27%) | 78 (33%) | 160 (24%) | <.0001 |
|  | Non-Hispanic White | 81814872 (62%) | 1398 (31%) | 92 (38%) | 332 (51%) |  |
|  | Non-Hispanic Black | 14783349 (11%) | 1038 (23%) | 43 (18%) | 81 (12%) |  |
|  | Other/ Multi-Racial | 12902446 (10%) | 902 (20%) | 26 (11%) | 81 (12%) |  |
| **Education** | < 12th grade | 14524629 (11%) | 764 (17%) | 63 (26%) | 110 (17%) | <.0001 |
|  | High school | 31911011 (24%) | 1089 (24%) | 67 (28%) | 177 (27%) |  |
|  | AA degree | 43807221 (33%) | 1488 (32%) | 84 (35%) | 257 (39%) |  |
|  | College graduate | 41605743 (32%) | 1242 (27%) | 25 (10%) | 110 (17%) |  |
| **Smoking** | Yes | 52729086 (40%) | 1353 (30%) | 185 (77%) | 507 (78%) | <.0001 |
|  | No | 79119517 (60%) | 3230 (70%) | 54 (23%) | 147 (22%) |  |
| **FPL** | <100% | 18801097 (14%) | 937 (20%) | 80 (33%) | 134 (20%) | <.0001 |
|  | 100 - 400% | 63541742 (48%) | 2475 (54%) | 123 (51%) | 342 (52%) |  |
|  | >400% | 49505765 (38%) | 1171 (26%) | 36 (15%) | 178 (27%) |  |
| **Insurance** | No insurance | 21385068 (16%) | 911 (20%) | 83 (35%) | 159 (24%) | <.0001 |
|  | Private insurance | 64221359 (49%) | 2027 (44%) | 57 (24%) | 252 (39%) |  |
|  | Medicaid/Medicare | 12821985 (10%) | 588 (13%) | 52 (22%) | 88 (13%) |  |
|  | Other insurance | 10881497 (8%) | 470 (10%) | 26 (11%) | 54 (8%) |  |
|  | Multiple insurance | 22538694 (17%) | 587 (13%) | 21 (9%) | 101 (15%) |  |
| **Employment** | Employed | 103710599 (79%) | 3369 (74%) | 159 (67%) | 479 (73%) | 0.0172 |
|  | Unemployed | 28138005 (21%) | 1214 (26%) | 80 (33%) | 175 (27%) |  |
| **Mean DMFT** |  | 7.86 - 8.50 | 7.86 | 8.46 | 9.82 | <.0001 |
| **Severe Caries** | No | 107491801 (82%) | 3742 (82%) | 175 (73%) | 488 (75%) | 0.028 |
|  | Yes | 24356803 (18%) | 841 (18%) | 64 (27%) | 166 (25%) |  |

Among current users, the majority (40%) were aged 18-30, while the largest proportion of former users (36%) were aged 51-59. Age distribution differed significantly across groups (p < 0.0001 The overall gender distribution was balanced; however, males predominated among both current (69%) and former users (58%), while non-users were primarily female (55%, p < 0.0001). Non-Hispanic whites constituted the largest racial group among both former (51%) and current users (38%) (p < 0.0001). Educational attainment was lowest among current users, with the highest proportion having less than a 12th-grade education and the lowest proportion being college graduates (p < 0.0001). Smoking was prevalent among current and former users, whereas most non-users were non-smokers (p < 0.0001). Current users had the highest proportion of participants with a family income below 100% of the federal poverty level (33%). Additionally, the percentage of uninsured individuals was highest (35%) among current users, with the lowest prevalence of private insurance (24%) compared to other groups (p < 0.0001).

Employment status varied significantly among the groups, with the highest unemployment observed among current users (33%, p = 0.017). Severe dental caries status also differed significantly between groups (p < 0.0001). Mean DMFT scores were highest among former users (9.82) and lowest among non-users (7.86) (Fig. 1). The prevalence of severe caries (DMFT > 13.99) was significantly higher among current users (27%) compared to non-users (18%, p = 0.028) (Table 1).

Tables 2 presents the distribution of severe caries, employment status, and covariates stratified by employment and oral health among the current users of cocaine, heroin, or methamphetamine. Bivariate analysis showed a significant association between unemployment and severe caries among both current (p=0.0008) and former users (p=0.0232). Among current users, insurance status and family income were significantly associated with employment status (p < 0.0001). Gender and education also showed significant associations with employment among current users (p < 0.05). While age, race, smoking, and insurance status were significantly associated with severe caries among former users, only age and family income showed significant associations among current users (p < 0.05).

**Table 2:** Table showing distribution of Severe caries, employment status, and covariates stratified by employment and oral health among the people who currently use cocaine/ heroin/ methamphetamine (n=239).

|  |  | Outcome: Employment |  |  | Exposure: Severe caries |  |  |
| --- | --- | --- | --- | --- | --- | --- | --- |
|  |  | Unemployed | Employed |  | No | Yes |  |
|  |  | N (%) | N (%) | p-value | N (%) | N (%) | p-value |
| Employment |  | 80 (33%) | 159 (67%) |  | 175 (73%) | 64 (27%) |  |
|  | Employed | 0 (0%) | 159 (100%) |  | 125 (71%) | 34 (53%) | 0.0008* |
|  | Unemployed | 80 (100%) | (0%) |  | 50 (29%) | 30 (47%) |  |
| Severe caries | No | 50 (63%) | 125 (79%) | 0.0008* | 0 (0%) | 64 (100%) |  |
|  | Yes | 30 (38%) | 34 (21%) |  | 175 (100%) | 0 (0%) |  |
| Age | 18-30 | 31 (39%) | 64 (40%) | 0.003* | 85 (49%) | 10 (16%) | <.0001* |
|  | 31-40 | 16 (20%) | 53 (33%) |  | 55 (31%) | 14 (22%) |  |
|  | 41-50 | 10 (13%) | 22 (14%) |  | 22 (13%) | 10 (16%) |  |
|  | 51-59 | 23 (29%) | 20 (13%) |  | 13 (7%) | 30 (47%) |  |
| Gender | Male | 48 (60%) | 118 (74%) | 0.0026* | 125 (71%) | 41 (64%) | 0.4588 |
|  | Female | 32 (40%) | 41 (26%) |  | 50 (29%) | 23 (36%) |  |
| Race | Mexican/Other Hispanic | 22 (28%) | 56 (35%) | 0.1599 | 64 (37%) | 14 (22%) | 0.452 |
|  | Non-Hispanic White | 33 (41%) | 59 (37%) |  | 65 (37%) | 27 (42%) |  |
|  | Non-Hispanic Black | 18 (23%) | 25 (16%) |  | 26 (15%) | 17 (27%) |  |
|  | Other/ Multi-Racial | 7 (9%) | 19 (12%) |  | 20 (11%) | 6 (9%) |  |
| Education | < 12th grade | 33 (41%) | 30 (19%) | 0.0218* | 46 (26%) | 17 (27%) | 0.2514 |
|  | High school | 21 (26%) | 46 (29%) |  | 50 (29%) | 17 (27%) |  |
|  | AA degree | 21 (26%) | 63 (40%) |  | 59 (34%) | 25 (39%) |  |
|  | College graduate | 5 (6%) | 20 (13%) |  | 20 (11%) | 5 (8%) |  |
| Smoking | Yes | 62 (78%) | 123 (77%) | 0.721 | 128 (73%) | 57 (89%) | 0.2808 |
|  | No | 18 (23%) | 36 (23%) |  | 47 (27%) | 7 (11%) |  |
| FPL | <100% | 34 (43%) | 46 (29%) | 0.0498* | 48 (27%) | 32 (50%) | 0.0188* |
|  | 100 - 400% | 39 (49%) | 84 (53%) |  | 96 (55%) | 27 (42%) |  |
|  | >400% | 7 (9%) | 29 (18%) |  | 31 (18%) | 5 (8%) |  |
| Insurance | No insurance | 27 (34%) | 56 (35%) | <.0001* | 56 (32%) | 27 (42%) | 0.064 |
|  | Private insurance | 11 (14%) | 46 (29%) |  | 48 (27%) | 9 (14%) |  |
|  | Medicaid/Medicare | 31 (39%) | 21 (13%) |  | 35 (20%) | 17 (27%) |  |
|  | Other insurnace | 5 (6%) | 21 (13%) |  | 20 (11%) | 6 (9%) |  |
|  | Multiple insurnace | 6 (8%) | 15 (9%) |  | 16 (9%) | 5 (8%) |  |

\* p-value < 0.05

The relationship between severe caries and employment was particularly strong among current substance users. Table 3 displays the unadjusted and adjusted odds ratios (OR), confidence intervals (CI), and p-values for the association between severe caries and employment status among those who used cocaine, heroin, or methamphetamine in the past year. Current users with severe caries were over three times more likely to be unemployed (OR = 3.23; 95% CI: 1.38-7.60; p = 0.008) compared to those without severe caries. After adjusting for insurance status (Model 2), the OR decreased to 2.59 but remained statistically significant. Further adjustments in Models 3 and 4 yielded similar ORs, and the associations continued to be significant. Among former users, severe caries was associated with a 1.7-fold increase in the likelihood of unemployment (OR = 1.7; 95% CI: 1.04-2.77; p = 0.036); however, this association lost significance after adjusting for insurance status and additional covariates.

**Table 3:** Crude and adjusted odds ratios of employment status associated with severe caries experience (DMFT >13.99) among current and former users of cocaine/heroin/methamphetamine after sequential adjustment for confounders.

|  | OR | 95% Confidence interval | P-value |
| --- | --- | --- | --- |
| **Progressive adjustment that is focused on DMFT > 13.99 among current users (n = 239)** |  |  |  |
| **Model 1:** Unadjusted | 3.23 | 1.38 - 7.60 | 0.0088* |
| **Model 2:** Insurance | 2.59 | 1.13 - 5.90 | 0.0253* |
| **Model 3:** Insurance, FPL, age, education | 2.64 | 1.15 - 6.07 | 0.0235* |
| **Model 4:** Race, age, insurance, gender, education, smoking, FPL | 2.57 | 1.13 - 5.86 | 0.0256* |

## Discussion

This study explored the relationship between dental caries and employment status among U.S. adults who have used cocaine, heroin, or methamphetamine. Utilizing data from the National Health and Nutrition Examination Survey (NHANES) from 2015 to 2018, we achieved our objectives. Our hypothesis—that severe dental caries would be associated with unemployment among current users of these illicit drugs— was supported. Current users with severe caries were 2.6 times more likely to be unemployed. However, among former users, no significant association was observed after adjusting for confounding variables. This is the first study to describe the association between dental caries and employment among a nationally representative sample of U.S. adults who use these substances, contextualizing the findings within the broader framework of social determinants of health (SDOH).

In our study, most current users were aged 18–30 years, aligning with the 2020 National Survey on Drug Use and Health, which reported the highest past-year illicit substance use among individuals aged 18–25 years (30). Non-Hispanic whites constituted the majority across population groups, but Mexican/Other Hispanics had the highest proportion among current users. We observed significant gender disparities in employment: males were significantly more likely to be employed than females among both current and former substance users. This finding supports Laudet’s study, which found that males were twice as likely to be employed among formerly polysubstance-dependent urban individuals (31). However, Miguel et al. reported no gender disparity among cocaine users in pooled clinical trial data (32).

Our study found that current substance users with severe dental caries were more likely to be unemployed (47%), uninsured (42%), and have a family income less than 100% of the federal poverty level (50%). This aligns with evidence that socioeconomic status significantly affects oral health access and outcomes (4). Limited education can influence substance abuse behaviors, affecting both oral health and employment opportunities (5).

Mean DMFT scores differed significantly among the three groups—current, former, and non-users—and were highest among former users, possibly due to the older age of this group. Notably, current users, despite being predominantly younger, had a higher prevalence of severe caries (27%) compared to former users (25%) and non-users (18%). This suggests a possible association between current drug use and increased dental caries, consistent with findings from Hegazi et al. (16) and Bahdila et al. (15), who reported increased dental caries among U.S. adults aged 30–64 years currently using cocaine or methamphetamine. Similar results were found by Cury et al. in Brazilian cocaine users (33). Our results also correspond with studies by Rommel et al. (34) and Shetty et al. (35), which found significantly higher xerostomia and DMFT among methamphetamine users. Methamphetamine reduces salivary flow and pH, altering dental plaque composition and volume, and substance users often consume more sugary, acidic beverages, exhibit poor oral hygiene, and lack adherence to treatment (36).

This is the first study to evaluate the association between dental caries and employment among people who use illicit drugs. Our findings are consistent with earlier studies reporting a strong correlation between employment status and oral health in people with substance use disorders (22–24). By utilizing NHANES data, we addressed a critical knowledge gap, providing insights that could inform policy and rehabilitation programs aiming to holistically improve the quality of life for illicit drug users.

The strong association between poor oral health and unemployment among illicit drug users suggests that dental care could be a crucial component in rehabilitation and employment programs. Incorporating comprehensive oral health interventions may facilitate the reintegration of this marginalized population into the workforce, improving their quality of life and decreasing societal burdens.

A major strength of this study is the use of NHANES data, which employs a three-stage stratified random sampling method and a large sample size, yielding nationally representative data. Dental examinations were performed by calibrated dentists, enhancing data validity and reliability. Controlling for a wide range of potential confounders strengthened the internal validity of our results. Including U.S. adults aged 18–60 years expanded the scope compared to previous studies limited to older NHANES cycles.

However, the study’s cross-sectional design limits the ability to ascertain temporality and causality between dental caries and employment. Self-reported substance use data may lead to underestimation due to recall and social desirability biases. The dental examination did not include radiographs, potentially underestimating dental caries prevalence. Some subgroups had fewer than 100 subjects, which may increase random error and widen confidence intervals. The categorization of current and former users based on use within or prior to the past year may not accurately capture the nuances of drug use patterns, possibly leading to underestimation of associations.

This study underscores the importance of oral health within the broader context of public health. We established a statistically significant association between severe dental caries and unemployment among adults who have used cocaine, heroin, or methamphetamine within the past year, even after accounting for insurance status. This emphasizes the intrinsic link between oral health and employability in this demographic. Our findings suggest that integrating dental professionals into multidisciplinary public health interventions could enhance the recovery journey of illicit drug users. Early screening and preventive dental services may not only help avert severe caries but also facilitate reentry into the workforce, boosting self-esteem and societal integration.

Given the limitations of our study, future research should focus on prospective investigations to conclusively establish causal links between oral health and employment outcomes among illicit drug users. Such research could significantly inform policy measures and public health strategies aimed at holistically improving the quality of life for individuals grappling with substance use issues. Additionally, interventions that operate across multiple levels of the social-ecological model may be more effective in addressing the complex interplay of factors influencing oral health and employment in this population.

## Conclusion

In summary, our findings reveal a significant association between severe dental caries and unemployment in adults reporting any cocaine, heroin, or methamphetamine use within the past year. While we were unable to measure the frequency or severity of drug use, the association observed in a nationally representative sample underscores the importance of oral health interventions for individuals with any history of illicit drug use. By addressing both the social determinants of health and the complex, interrelated effects of substance use on oral health, policymakers and public health practitioners may enhance employability and overall quality of life for at-risk populations. Future research should incorporate longitudinal designs and granular substance use measures to determine how differing levels of use influence these outcomes and to inform more specialized interventions.

## Data Availability

https://www.cdc.gov/nchs/nhanes/index.html

